# “This is really like waiting for war and this is not good” – Intertwining between pandemic experiences, and the development of professional action of healthcare professionals in critical care at the beginning of the COVID-19 pandemic in Germany: a qualitative study

**DOI:** 10.1101/2021.01.29.21250626

**Authors:** Madlen Hörold, Karl Philipp Drewitz, Vreni Brunnthaler, Julia Piel, Magdalena Rohr, Ilona Hrudey, Claudia Hasenpusch, Angela Ulrich, Niklas Otto, Susanne Brandstetter, Christian Apfelbacher

## Abstract

Healthcare professionals (HCPs) are facing remarkable challenges in their daily work since the outbreak of the COVID-19 pandemic. Being well prepared is crucial for dealing with such a pandemic. The aim of our study was to explore HCPs’ subjective perspectives on their professional action and coping strategies in critical care during the preparation and coping phase after the outbreak of the COVID-19 pandemic in Germany.

Together with HCPs working in critical care, we collaboratively designed an interview study based on an ethnomethodological approach. We performed semi-structured qualitative interviews via telephone or video call and analysed the data based on grounded theory.

Our research interest was focused on HCPs (qualified nurses, physicians, medical students) working in critical care during the first wave of the COVID-19 pandemic in Germany between April and July 2020.

Our sample consisted of 39 HCPs (19 nurses, 17 physicians, three medical students, 18/39 female) from ten German federal states. All participants were involved in the acute care of COVID-19 infected patients in hospitals and had a mean professional experience of 14.8±10.1 years, 15 participants held a management position (e.g. senior physician or head nurse). We recruited participants via personal contacts and snowballing.

Initial and focused coding resulted in seven categories: Creating structural measures, handling operational changes, dealing with personal protective equipment, building up knowledge and skills, managing information, perceiving peer support and experiencing emotions.

Professional action and subjectively perceived preparedness (professional and emotional) interacted with each other. Their interrelation was not static, but rather dynamic and ambiguous according to the situation. The findings of our study can be beneficial in developing guidelines, policy interventions or personnel and work practice strategies.

## Introduction/ Background

Since the outbreak of the Coronavirus Disease (COVID)-19 pandemic, health care systems across the world have been facing unprecedented challenges in continuously re-organizing (intensive) care. In the beginning, strategies for preparing for rapidly changing situations of care were accompanied by substantial uncertainty [1–3]. The German health care system has first been confronted with the novel severe acute respiratory syndrome coronavirus type 2 (SARS-CoV-2) causing COVID-19 in January 2020 [4].

The extent to which healthcare professionals (HCPs) are prepared or can be prepared for unforeseeable, dynamic changes and their impact on the care situation are decisive in determining whether comprehensive care can be provided for critically ill COVID-19 patients. Previous studies from emerging infectious diseases such as the SARS (2002–2004) or Ebola (2014-2016) outbreaks show consequences for nurses like loneliness, fatigue and sleep disorders or even posttraumatic stress disorder due to the extremely high workload and additional social isolation caused by quarantine measures [5–7]. In addition, the risk of self-infection causes anxiety among HCPs including the fear of infecting their next-of-kin and of the increased workload due to infected and thus non-working colleagues [8–12].

Previous studies focused on psychological consequences of the pandemic. In a viewpoint, Shanafelt et al. [13] e.g. aimed to understand and address sources of anxiety among HCPs during the COVID-19 pandemic and described eight sources of anxiety, including access to appropriate personal protective equipment (PPE) and lack of access to up-to-date information and communication. Sun et al. [14] interviewed 20 nurses of the first affiliated hospital of Henan University about their psychological state and identified four topics: the presence of negative emotions at the beginning, the simultaneous or gradual appearance of positive emotions, coping mechanisms and growth under stress. In another qualitative study, Liu Yu et al. [15] described four key issues in the professional context of the respondents: Challenges and dangers, pressure due to fear of infection and high workload, strong sense of duty and a rational understanding of the epidemic. In a further study by Liu Qian et al. [16], the authors reported on three emerging issues: Responsibility for patients, challenges in the workplace and resilience in the midst of these challenges. Authors of a recent rapid review [17] including 44 studies on evidence about the psychological impact of epidemic or pandemic outbreaks on HCPs recommended that coping strategies for HCPs should be assessed and promoted as well as that sufficient PPE should be provided in order to “mitigate […] negative psychological responses of” HCPs [17].

It is thus of essential importance to identify the strategies that HCPs are developing within their institutional environment to maintain the quality of professional care, to find out what decisions are made and which processes are initiated to re-adjust workflows and to provide appropriate care to patients under the conditions of the pandemic. To address this research gap, we set out to explore HCPs’ subjective perspectives on professional action and coping strategies in acute care during the preparation and coping phase after the outbreak of the COVID-19 pandemic in Germany from March to July 2020. The main research interest of our study was to expose implicit principles that structure social practice and interaction of HCPs [18] in an exceptional situation which is characterized by a high level of change.

## Material and Methods

### Ethical considerations, data protection and privacy

We received ethical approval for our research from the institutional review boards of the University of Magdeburg (51/20) as well as the University of Regensburg (20-1771-101) before we performed the first interview. All study activities were conducted in accordance with the declaration of Helsinki [19] and in compliance with the relevant legal regulations. Each participant received and signed a declaration of consent before we made an appointment for the respective interview. We interviewed participants individually via telephone or an appropriate video conferencing system and recorded the interview in an audio format compliant with the General Data Protection Regulation, GDPR [20]. We assured confidentiality by assigning a five-digit number for each participant. To make the results easier to read, we generated pseudonyms beyond the non-speaking numbers. These do not allow any conclusions to be drawn about the true identity of the study participants. To further fulfil GDPR compliance, we set up a trusteeship.

### Study design and participants

The study is based on Garfinkel’s ethnomethodological approach [18, 21] and considers everyday professional action under the circumstances of the first phase of the COVID-19 pandemic in German hospitals. In order to explore how HCPs adapt their professional actions to build up “normality”, we contacted 129 hospitals or individual HCPs throughout Germany (across all 16 federal states) by different media (e-mail, telephone, professional networks, distribution of a flyer) and personal contacts via snowball sampling [22] between end of March and mid July 2020. We addressed persons from several health care professions e.g. physicians, academically qualified nursing staff and medical students in German hospitals, who were involved in the clinical acute care of COVID-19 patients requiring intensive care or monitoring. Two contacted individuals actively declined to participate in the study at the first point of contact without specific reasons, while two asked to be contacted only after the pandemic. 39 agreed to participate. All other contacts did not respond or did not get back in touch after initial communication. There was no drop-out of participants between recruitment and the actual interview. To gather rich data [23], we aimed for a heterogeneous sample both in terms of individual characteristics (e.g. work experience, gender, social and ethnic origin, educational background) and the professional environment (including the level of care provided by the hospital). Repeat interviews are carried out and will be published at a later stage.

### Involvement of patients/participants and public

Patients or the public were not involved. In the beginning of the pandemic outbreak, our research team was approached by HCPs with the idea for this study. We developed the study protocol collaboratively and were supported by the initiators during the recruitment process (snowballing). All participants were informed in advance about our publication strategy, which is in line with our study protocol.

### Data collection

We developed a semi-structured thematic interview topic guide (see supplementary information) based on the most relevant emerging issues and discussion points of the federal government, federal states governments, global research activities and public opinion regarding the challenges of the pandemic. This semi-structured approach supported the different interviewers in keeping the focus on the research interest and, if necessary, in responding adequately to specific events in the interview situation. Participants were initially asked about their personal experiences and feelings regarding the preparations of the institution they worked in. For this purpose, we used a narrative stimulus based on a method of Fritz Schütze [24] at the beginning of the interview: *“I would like to ask you to tell us how you perceived the phase of preparation for the COVID-19 pandemic in your professional context and how you experienced it. First of all, you can take as much time as you like and speak about whatever comes to your mind*.*”* The open beginning of the interview gave the participants the opportunity to set their own relevance, to express the subjective meaning of the topic and to reflect on what they have experienced. They were encouraged beforehand to use an artefact (e.g. photographs, newspaper articles, objects, etc.) that reflected the current work situation from their perspective.

All interviewers (n=7) were female with a varying degree of experience in conducting qualitative research. Four had prior experience in healthcare (one nurse, one radiographer, one psychologist, one physiotherapist). The coding and interpretation team consisted of two further male researchers and one female researcher, who also had prior experience in conducting qualitative interview studies. The entire team was composed of one professor (CA), two senior lecturers/postdoc researchers (MH, SB), three research fellows/PhD-candidates with master degree (PD, JP, MR), three junior scientists with master degree (VB, IH, NO) and two junior scientists with bachelor degree (CH, AU). Study participants belonging to the researchers’ personal networks were assigned to other interviewers they did not know. Participants were informed about the professional background of the interviewers, the aim of the research and again about the relevant data protection and privacy issues. In order to contextualize the insights gained from the interviews, socio-demographic data, information on professional biography, the current situation within the current participants’ workplace and various aspects of the interview situation i.e. atmosphere and interaction, were collected as data of secondary order. Some participants had a short preliminary interview as part of the scheduling process. Two interviews were conducted as pilot interviews but included in the sample.

### Data processing, analysis and reporting

The audio recordings were transcribed verbatim and pseudonymised. All information allowing the identification of the interview partner was removed. We analysed the interviews in a collaborative interpretation process based on grounded theory [23, 25]. The quality and methodically controlled procedure of inductive data analysis was achieved by communicative validation within the research group through regular meetings via a video conferencing system [26]. Data was categorised into segments in order to achieve analytical accounting and build up emergent theoretical concepts [23]. During the computer-assisted (performed with either Atlas.ti or MAXQDA) data analysis, we switched between line-by-line, in-vivo, incident-to-incident and focused coding, guided by Charmaz [23]. We documented the results of the group analysis in tabular form. For collaborative interpretation, we used Confluence by Atlassian. The focused codes were collected with systematic memos accompanying the research process in a table in order to continue define conceptual categories by further coding steps [23]. We initially coded in German and then translated into English. Reporting of this study is based on the Consolidated Criteria for Reporting Qualitative Studies (COREQ) Checklist [27]. Respondent validation was not considered.

## Findings

### Study sample

The sample consisted of 39 HCPs: 19 nurses, 17 physicians and three medical students from ten German federal states (see Figure 1). 46% were female. All participants were involved in the acute care of COVID-19 infected patients in hospitals and had a mean professional experience of 14.8±10.1 years, 42% held a management position (e.g., senior physician or head nurse). They entered COVID-19 care at different times, from mid-March to the end of May. The interviews were conducted between April 6th and July 13th and lasted about 12-66 minutes (mean 34.8 ±11.8 minutes). About half (46%) was working in maximum care hospitals (e.g., university hospitals). Further details on the characteristics of the participants are shown in Figure 1.

**Figure 1:**
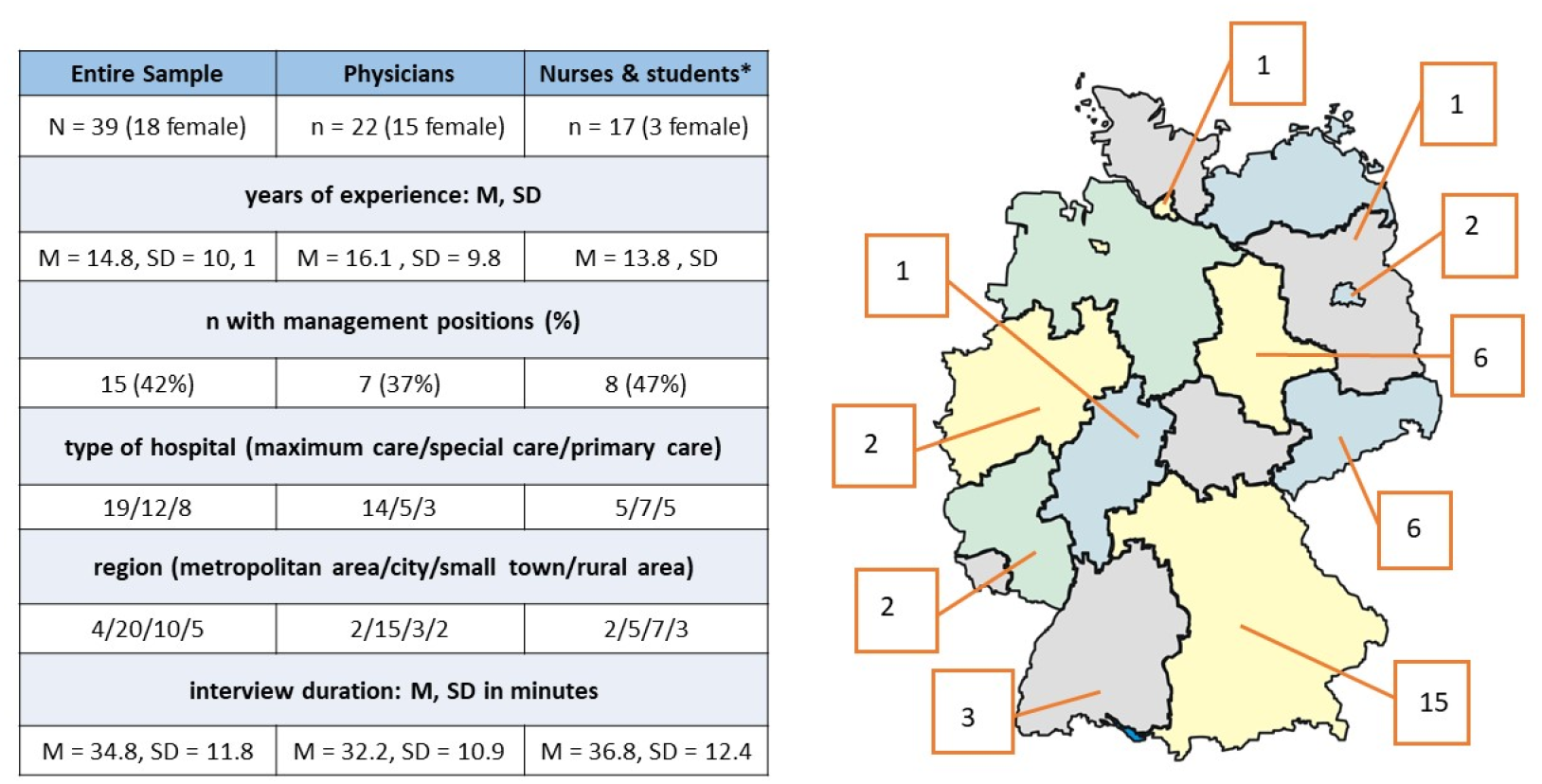
On the left side: Characteristics of the study sample; on the right side: Regional distribution of participants for each German federal state (Bundesland) *) 3 medical students, who had nursing experience before and volunteered on the respective ICU during the pandemic

### Categories and concepts

A focus of questioning the data was on thematic redundancies the participants made, and what theories, motives, evaluations, justifications and further explanations as well as symbolic markers were referred to frame described actions. In this context, we already became aware of the significance of metaphorical terms that invoke martial or catastrophic scenarios during initial coding.

Initial and focused coding resulted in seven inductively created categories each with subordinate concepts on experiences and the development of professional actions and coping strategies of HCPs in acute care:

- ***Creating structural measures***,
- ***Handling operational changes***,
- ***Dealing with personal protective equipment***,
- ***Building up knowledge and skills***,
- ***Managing information***,
- ***Perceiving peer support* and**
- ***Experiencing emotions***.

The first concept within the category ***“Creating structural measures”*** was *searching and finding solutions*. In the hospitals, multidisciplinary crisis teams or task forces were set up as an extension of the hospital management, which centrally determined the measures for the respective facilities (based on legal requirements). The task forces met daily or weekly. Their decisions had a strong impact on the structures and operational processes. The second concept was *building up COVID-19 units*. The participants reported that different organizational measures have been undertaken to create capacities for a possibly large number of COVID-19 infected persons. Thus, intensive care units (ICU) were extended (partly also by structural measures), regular wards were closed, merged, medically rededicated or equipped with other medical technology. As a third concept, we elaborated *managing human resources*. In order to increase personnel capacities, employees were reallocated, qualified or recruited (with and without previous medical/nursing experience). In addition, the support of volunteers was requested. Frontline workers received support through, for example, the offer for talks with chaplains or a crisis intervention team, free drinks and food, massages or even staff supervising the correct donning and doffing.

The second category ***“Handling operational changes”*** consisted of three concepts: *Implementing new workflows, balancing voluntary work and duty*, and *task sharing and changing responsibilities*. The participants reported about implementing new workflows in their units through operational changes. For example, access to the departments was restricted or zoned within the COVID-19 units (similar to a traffic light system). Routines got lost, workflows were adapted, and support services offered. Locking in and out of patients’ rooms was reduced to a minimum to protect against infection. Accordingly, all activities in the patients’ room and the required equipment had to be well prepared. The *balancing voluntary work and duty* concept was an expression of an ongoing negotiation process of the management. On the one hand, employees (physicians and nurses) who volunteered to care for COVID-19 patients staffed units. On the other hand, employees were confronted with the care of COVID-19 patients through the reassignment of areas. In particular, staff with personal risk profiles for severe progression of COVID-19 if infected were looking for ways out of this service commitment, sometimes solutions were found. The concepts *task sharing and changing responsibilities* described changes in direct patient care. The participants talked about the support of colleagues without ICU skills, auxiliaries and service staff and the associated assignment/allocation of tasks, as well as changes in rostering (floater, digitalization). Individual physicians and nurses described a change of roles in the patient room: Some doctors took over nursing tasks and vice versa. Some nurses reported that ward rounds were carried out without doctors seeing the patients.

The third category ***“Dealing with personal protective equipment”*** *(PPE)* consisted of three concepts relating to organisational, operational and individual aspects. We called the first concept *rationing*. The participants described a deficiency and rationing of PPE. Due to theft, the equipment had to be stored in locked rooms and was only handed out personally to each employee per shift. This also involved the implementation of new procedures. The second concept was *opening up creative procurement channels*. Participants reported many different efforts by hospitals; such as purchasing in the hardware store, using the private neighbour’s 3D printer. High prices were paid to medical device companies to provide protective equipment to employees. With regard to *lowering quality standards* as the third concept, participants described how they suddenly had to use respirators and protective gowns more than once, despite previous hygiene rules.

The fourth category ***“Building up knowledge and skills”***, consisted of the three concepts *donning and doffing, training ICU skills* and *being instructed in new devices*. Participants defined *donning and doffing* as an important training measure in infection control. Teaching material such as video sequences was provided, and techniques were practiced under supervision. In addition, participants told of (short) training courses to build up ICU skills. Employees without or with little ICU experience were prepared for a possible assignment. There were differences in training contents and scope. However, the focus was mainly on equipment, ventilation and monitoring. In order to ensure the care of a potentially large number of critically ill patients, all available equipment, especially (home-) respiration, injection pumps and dialysis machines, was (re)activated and partly newly purchased. Accordingly, employees were instructed in how to work with these devices.

The fifth category ***“Managing information”*** consisted of four concepts: *Being confronted with flood of information with limited validity, exploring new digital world, talking and listening* and *being confronted with “Corona” everywhere*. Within the first concept, the participants described their efforts in collecting information and difficulties in managing its large amount (by handout, e-mail and telephone). Within a very short time, management revised work instructions and process descriptions. The second concept was called *exploring new digital world*. The participants talked about an increasing use of the intranet, about video messages and instructional videos up to the development of apps for rostering. Especially supervisors described the importance of *talking and listening* (third concept) during this time. They mentioned the importance of listening to employees’ needs and concerns, answering their questions and explaining current measures. The fourth concept was called *being confronted with “Corona” everywhere*. The participants described the COVID-19 pandemic as an omnipresent phenomenon that accompanied their everyday communication both in the professional and the private environment.

The sixth category covered the field of ***“Perceiving peer support”***. In dealing with professional changes participants reported perceiving peer support. This category consisted of two concepts: *Feeling supported by family* and *friends and feeling collegiality and teamwork. The participants* mentioned family and friends as an important resource. They listened, were simply there or took care of the children. Furthermore, with regard to the second concept *feeling collegiality and teamwork*, the participants talked about experiencing support from colleagues and about working in a (multi-professional) team. They described changes in teams through new colleagues and structures. The team spirit seemed to have an impact on the experience of support. Successful teamwork was perceived as a great value.

The seventh category, ***“Experiencing emotions”***, described the different and sometimes dynamic emotional world of the participants, which we represented in the concepts of *having negative emotions, being proud and satisfied* and *being “Corona heroes”*. The participants in particular, expressed stress during work, although often fewer patients were treated on ICU than before the pandemic. They also spoke about worries, fears, frustration, anger and doubts, isolation (concept of *having negative emotions*). Worries and fears were often related to the lack of PPE and information, to the perceived uncertainty about the course of the pandemic, the experience of lack of structure and the assessment of one’s own resilience and the possibility of family support. The participants mentioned frustration and anger, especially in relation to the lack/rationing of PPE and staff shortages. Some participants felt that superiors did not sufficiently inform about strategic decisions nor supported them in their operational problems. Nurses and physicians in a management position expressed doubts about being capable to develop good and sustainable solutions and lead the hospital through the pandemic. All these negative feelings were also related to the experience of psychological burden. In contrast, participants reported also *being proud and satisfied* (second concept) of their own work, the possibility to help. They gained self-affirmation through their commitment. This seemed to be the driving force behind the motivation to work. We particularly identified motivated participants as “*Corona Heroes*”. They described the COVID-19 pandemic as eventisation and possibility of (public) attention leading to an increase in motivation and engagement. The ***“Experiencing emotions”*** category with its concepts overlapped with most of the other categories: Emotions were influenced by an individual’s behaviour as well as by the behaviour of others. Table 1 presents the inductive categories with respective concepts in an aggregated overview. Figure 2 contextualises the categories and relates them to each other. It is part of the coding and interpretation process to elaborate the central phenomenon and to transfer the structure of the emerged categories into a theory.

**Table 1:**
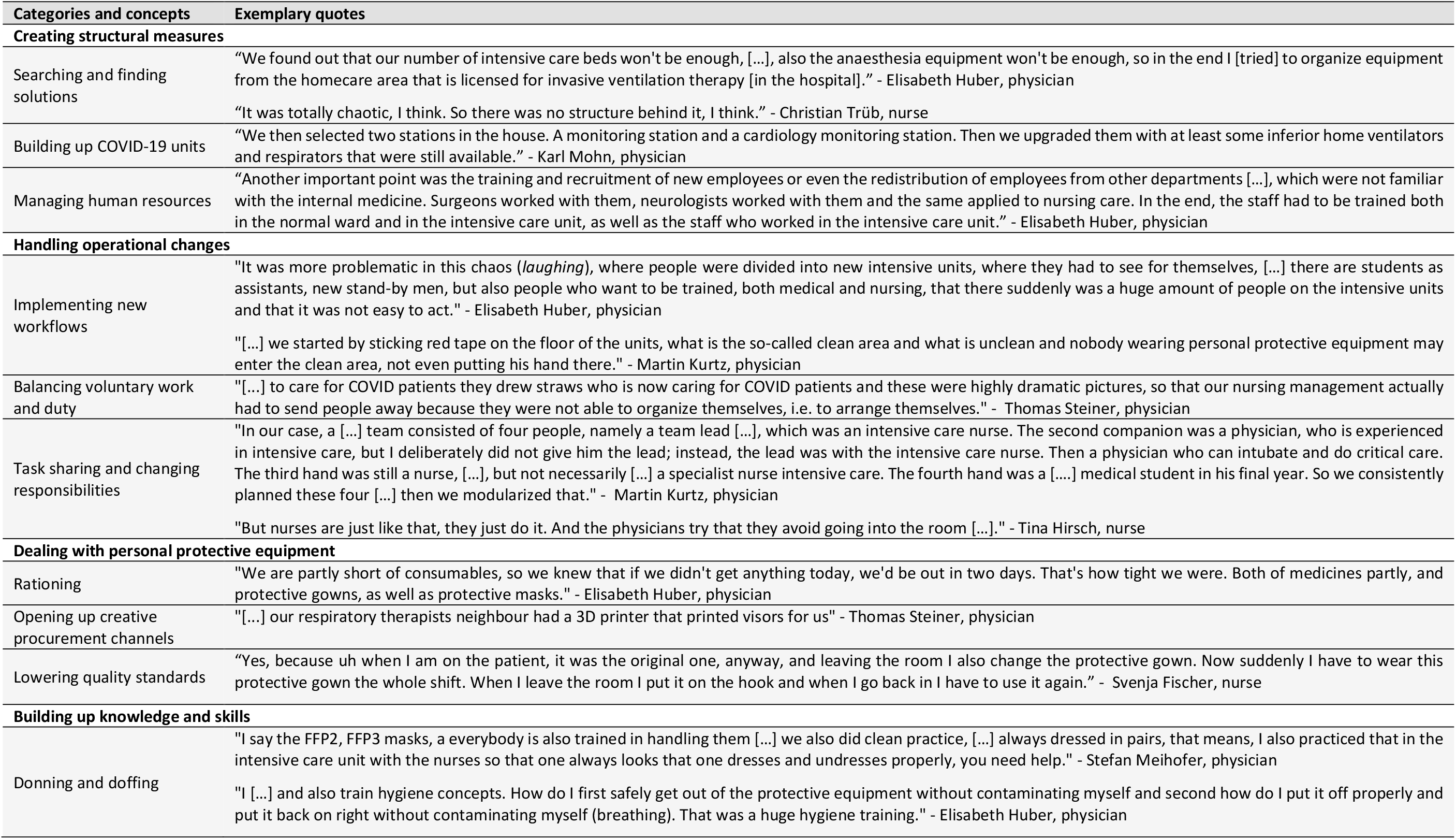
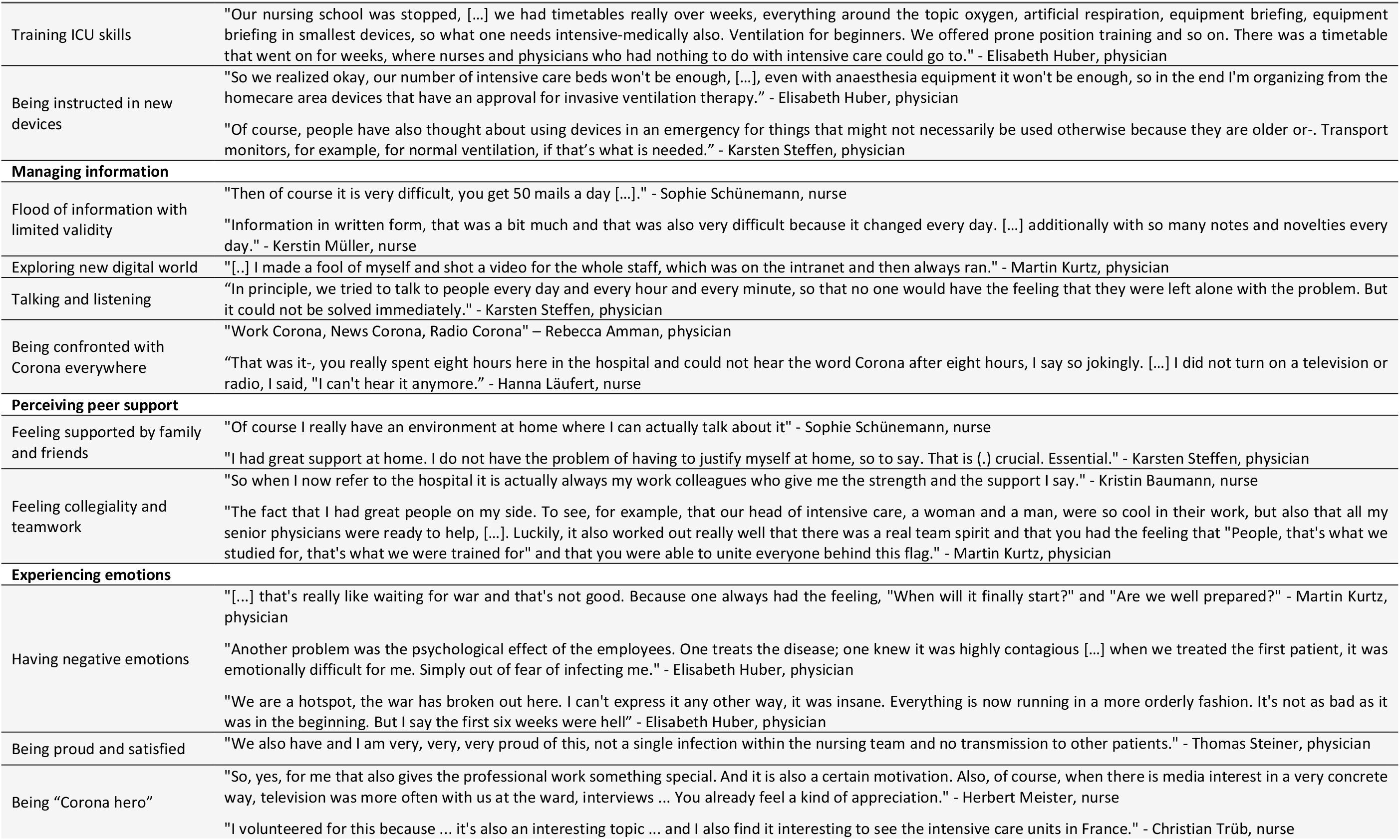
Aggregated overview of inductive categories with respective concepts and exemplary quotes

**Figure 2.**
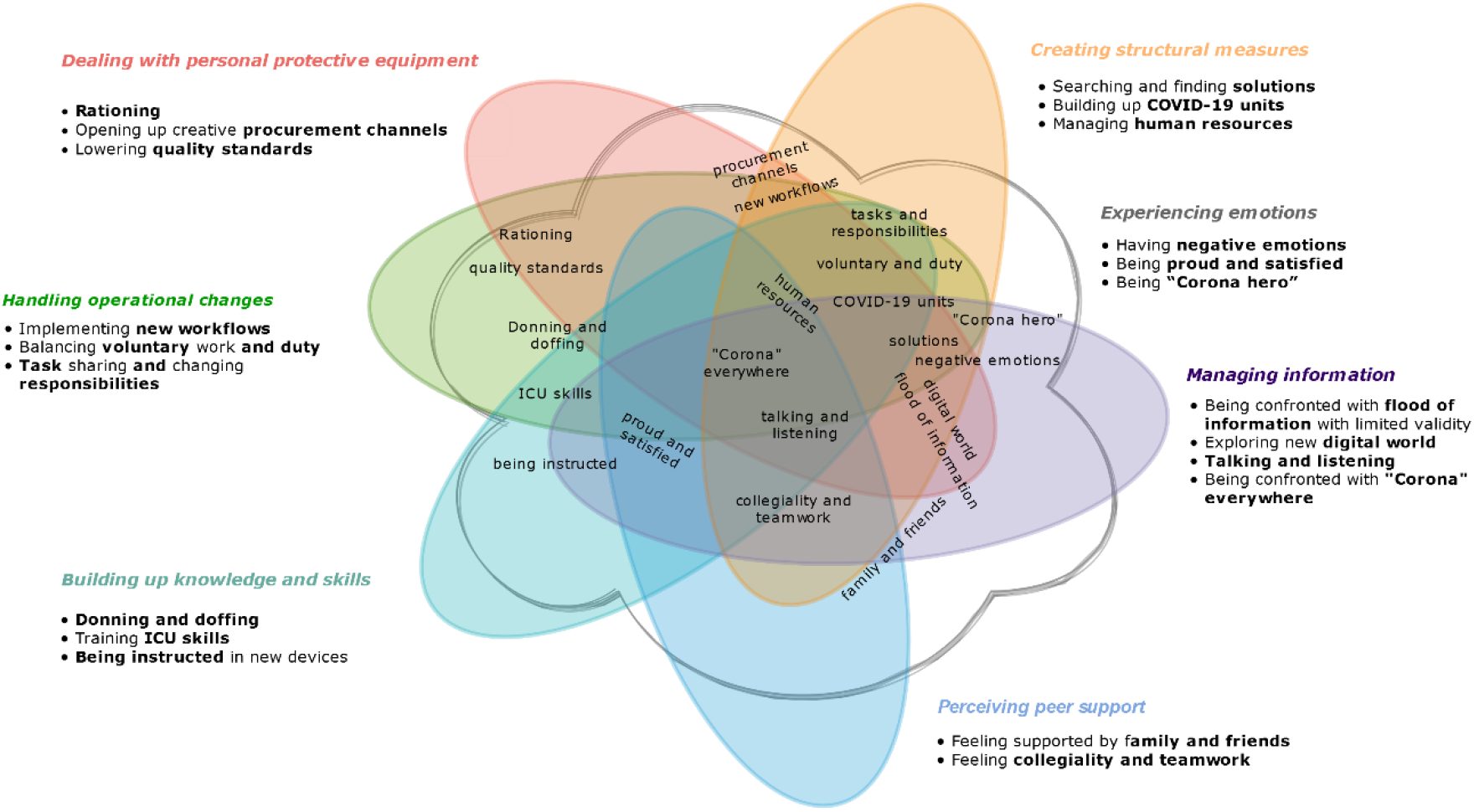
Situational map of intertwining of different phenomena/processes discovered during interpretation

The identified categories were closely interwoven; their interrelation was not static, but rather dynamic according to the situation. We were able to map trajectories between the categories in a multidimensional framework (see Figure 2). This first heuristic modelling of the categories gives a preview of a theoretical model that will be continued [23]. It represents the interpretation of the perspective of the respondents and summarizes the processes in the field. Based on the multidimensional arrangement of the categories and concepts, we kaleidoscopically arranged intersections, which suggest social interactions on various levels and provide indications of the emergence of “agency” from the perspective of the HCPs under the dynamic situation.

In the following we will explain the intertwining using examples. At first, the concept of having *negative emotions* and the associated stressful experiences. The participants talked about nerve-racking situations in connection with direct patient care, especially through PPE and adaptation to new operational measures. Furthermore, stressful feelings were discussed in relation with managing information, and the perceived lack of leadership (*→ Creating structural measures*).

(The concept-) ***Lowering quality standards*** (in the category *dealing with PPE*) resulted in new workflows and was thus part of the operational measures in hospitals. Participants reported new, lower quality standards regarding PPE, especially through rationing. This also triggered (negative) emotions among the participants. They expressed worries, fears and anger.

The concept ***human resources*** in the *Creating structural measures* category has intertwining with three other categories. The participants reported that employees were redistributed within the hospital, which created new workflows (handling operational changes). The management also recruited staff (with and without previous medical/nursing experience) and initiated training for existing and new employees with the necessary knowledge and skills to treat COVID-19 patients. In this context, there are interactions with emotions as well. Participants declared concerns about a perceived lack of staff and the loss of routines.

The concept of ***collegiality and team*** belongs to the *Perceiving peer Support* category. This concept also has intertwining to other categories/concepts. As a consequence of operational measures, changes in team structures and cooperation occurred. The teams provided support in managing information and dealing with PPE, through the possibility of talking and listening and in handling new workflows. The participants also reported on effects on their emotions: On the one hand, teamwork was associated with positive emotions. On the other hand, dysfunctional teams led to negative emotions among participants.

## Discussion

We examined how HCPs constructed the frame of professional action during the preparation and coping phase for the care of COVID-19 patients in Germany. The focus was on investigating HCPs’ specific experiences and associated actions. Seven main categories were identified from participants’ accounts: *Creating structural measures, handling operational changes, dealing with personal protective equipment, building up knowledge and skills, managing information, perceiving peer support* and *experiencing emotions*.

We found that the identified categories and concepts in our data were highly dynamic, as well as the resulting cause-and-effect forces, both contributing to changes in professional actions. Adopting a systemic perspective, we showed that processes in critical care always need to be considered multi-dimensional and that changes on one level always have an impact on other areas. Respondents used the interview as an opportunity for personal voice. Part of the sample complained about inadequate working conditions, which were intensified with the pandemic. Understanding the experiences gained by HCPs can lead to recommendations for action.

During a time of intense workload for people working in acute care we succeeded in recruiting 39 HCPs, from different regions in Germany, from high and low volume hospitals, before, during or after the care for the first wave of COVID-19 patients. Both nurses and physicians were included, reflecting everyday work in hospitals, but the sample consisted of mainly white German-speaking persons. Increasingly people with a migrant background are working in the healthcare sector and their perspectives may be different. Further, our sampling strategy was likely to identify participants who are highly motivated or particularly concerned about the pandemic. Data collection and analysis was organised in an ongoing circle allowing for maximum openness and for adjusting the design in the course of data collection and data analysis [23]. As interviews were conducted using telephone or video conferencing systems, it was sometimes difficult to build a trusting relationship with participants, since non-verbal cues could not be obtained. The subsequent translation of the research results and quotations carries the risk of losing or alienating the meaning [28]. Gender-specific dynamics in the interviews cannot be excluded, since only women conducted all interviews [29]. The varying degree of experience in conducting qualitative research or prior work experience in healthcare (four/seven interviewers) might have influenced the data collection and analysis, too.

Some of our study findings relate to the results of other studies conducted in the context of the COVID-19 pandemic. The key category - *experiencing emotions* - showed how the pandemic and the associated work-related processes/phenomena triggered emotions in HCPs, or how emotions influenced the experience of work-related processes/phenomena. Experiencing emotions interacted with all other categories; their interrelation was not static, but rather dynamic and ambiguous according to the situation. This is in line with Sun et al. [14], who also indicated that positive emotions occurred simultaneously with negative emotions. They discovered the phenomenon “growth under pressure”, which included increased affection and gratefulness, development of professional responsibility, and self-reflection [14].

Our findings regarding PPE and (psychosocial) support are consistent with previous published studies [14, 17, 30, 31]. Vindrola-Padros et al. [32] reported about limited PPE and lack of routine testing and the relation to anxiety and distress in HCPs. When PPE was available, incorrect size and overheating made daily work difficult. Lack of training for redeployed staff and the failure to consider the skills of redeployed staff for new areas were identified as serious challenges. Positive aspects of daily work reported by HCPs included solidarity between colleagues, the establishment of support structures and feeling valued by society.

Authors of a recent rapid review [17] recommended that coping strategies for HCPs should be assessed and promoted as well as that sufficient PPE should be provided in order to “mitigate […] negative psychological responses of” HCPs [17]. Adjustments of hospital infrastructure to COVID-19 (e.g. sufficient staff, keeping teams and working schedules stable) could support HCPs [31].

Preparing HCPs and encouraging (organisational) coping strategies seems to be very important in order to prevent anxiety, uncertainty, conflicts within working teams and long-term negative health consequences. Experiences of HCPs during the first SARS outbreak have shown the relevance of professional as well as emotional preparedness of HCPs with regard to coping strategies in a new care situation [14]. Employers of HCPs should be/become aware of the dynamics and complexity of the system and support coping and the development of professional actions through appropriate measures, e.g. regular and intensive training [16]. Conditions of insufficient human/material resources or lacking life-support measures, leading to end-of-life decisions [33–35] should be avoided. Our study highlights the importance of complexity of critical care and deriving situation-specific measures. It also demonstrates that positive emotions among employees are motivating; challenging circumstances can provide precious experiences and important insights - especially regarding teamwork, workflow and responsibilities. Therefore, it would be beneficial to consider the described experiences of the HCPs as learning processes and to feed them back into practice. Furthermore, our results can be beneficial in developing guidelines regarding infrastructure, policy interventions or personnel and work practice strategies.

By now, hospitals in Germany are challenged by the second wave of COVID-19 patients. We do not know whether experiences from the first wave as expressed by our study participants were used for improving the working conditions and quality of care and how HCPs evaluated their own and their hospitals actions during the pandemic retrospectively. Future studies which adopt a longitudinal perspective are necessary.

## Supporting information

coreq_checklist

## Data Availability

Data can be obtained from the corresponding author upon reasonable request.

## Declarations

CA is principal investigator of a study which develops and pilot tests an intensive care follow-up clinic. Further, he is spokesperson of the working group “Intensive care and critical illness” of the German Network Health Services Research, and member of the Scientific Advisory Board of the Eric project. All other authors declare no conflict of interest. All authors read the final version of the manuscript and approved its submission for publication. The corresponding author attests that all listed authors meet authorship criteria and that no others meeting the criteria have been omitted.

### Transparency declaration

The lead authors (the manuscript’s guarantors) affirm that the manuscript is an honest, accurate, and transparent account of the study being reported. No important aspects of the study have been omitted. Discrepancies from the study as have been explained.

### Dissemination declaration

Study participants will receive an overview of preliminary results after the follow-up interview in the form of a conference abstract. Further, the submission of this article will be followed by the dissemination of the study findings in plain language to the relevant professional associations.

### Author contributions (CRediT – Contributor Roles Taxonomy)

Madlen Hörold (formal analysis, investigation, methodology, validation, visualisation, writing/original draft, writing/review&editing), Karl Philipp Drewitz (conceptualisation, data curation, formal analysis, methodology, project administration, supervision, validation, visualisation, writing/original draft, writing/review&editing), Vreni Brunnthaler (data curation, formal analysis, investigation, methodology, project administration, validation, visualisation, writing/original draft, writing/review&editing), Julia Piel (formal analysis, investigation, methodology, validation, visualisation, writing/original draft, writing/review&editing), Magdalena Rohr (conceptualisation, data curation, formal analysis, methodology, project administration, validation, visualisation, writing/review&editing), Ilona Hrudey (formal analysis, investigation, validation, writing/review&editing), Claudia Hasenpusch (formal analysis, investigation, validation, writing/review&editing), Angela Ulrich (formal analysis, investigation, validation, writing/review&editing), Niklas Otto (formal analysis, investigation, validation, writing/review&editing), Susanne Brandstetter (formal analysis, methodology, supervision, writing/review&editing), Christian Apfelbacher (conceptualisation, funding acquisition, methodology, project administration, supervision, writing/review&editing)

### Data sharing

Data can be obtained from the corresponding author upon reasonable request.

## Acknowledgements

We are indebted to Maximilian Malfertheiner for initial discussions which led to the planning of the study, and to Julika Loss for supporting the study through additional intramural funding. Further, we are grateful to Stefanie March for advice in the planning of the study, to Heike Hupach for supporting the recruitment of potential study participants and to Christoph Damm for his input in the interpretation phase of the study. We thank Johannes Bernarding and Markus Plaumann for their advice and establishment of a trusteeship as part of our study. We would like to thank all the interview participants for their time and their openness to talk to us.

## Supplementary information

COREQ Checklist – uploaded separately

