## Supplementary material for "“This is really like waiting for war and this is not good” – Intertwining between pandemic experiences, and the development of professional action of healthcare professionals in critical care at the beginning of the COVID-19 pandemic in Germany: a qualitative study": coreq_checklist

| No | Item | Guide questions/description | Answers | Referred in manuscript on |
|---|---|---|---|---|
| **Domain 1: Research team and reflexivity** | | | | |
| Personal Characteristics | | | ... | |
| 1. | Interviewer/facilitator | Which author/s conducted the interview or focus group? | MH, MR, VB, IH, AU, JP, CH | Page 4 |
| 2. | Credentials | What were the researcher's credentials? *E.g. PhD, MD* | PhD, MPH, MSc, MSc, B.A., M.A., BSc (highest degree, respectively) | Page 4 |
| 3. | Occupation | What was their occupation at the time of the study? | senior lecturer, research fellow, research fellow, junior scientist, junior scientist, research fellow, junior scientist | Page 4 |
| 4. | Gender | Was the researcher male or female? | All interviewers were female, interpretation was done by eight female and two male researchers. | Page 4 |
| 5. | Experience and training | What experience or training did the researcher have? | All interviewers were experienced in conducting qualitative interviews for at least one prior research project. | Page 4 |
| Relationship with participants | | | | |
| 6. | Relationship established | Was a relationship established prior to study commencement? | Some interviewees were personally known by some of the researchers. We tried to avoid any bias by assigning those participants to other researcher.<br>Some participants had a short preliminary interview as part of the scheduling process. | page 4 |
| 7. | Participant knowledge of the interviewer | What did the participants know about the researcher? e.*g. personal goals, reasons for doing the research* | Participants were informed about the professional background of the interviewer and the aim of the research. | page 4 |
| 8. | Interviewer characteristics | What characteristics were reported about the interviewer/facilitator? e.g. *Bias, assumptions, reasons and interests in the research topic* | Participants were informed about the professional background of the interviewer. Prior work experience in healthcare (four/seven interviewers) | page 4 and page 12 |

| No | Item | Guide questions/description | Answers | Referred in manuscript on |
|---|---|---|---|---|
| | | | might have influenced the data collection. | |
| **Domain 2: study design** | | | | |
| Theoretical framework | | | | |
| 9. | Methodological orientation and Theory | What methodological orientation was stated to underpin the study? *e.g. grounded theory, discourse analysis, ethnography, phenomenology, content analysis* | The study is based on Garfinkel's ethnomethodological approach.<br>Coding was guided by grounded theory (approach from Charmaz). | page 3<br>page 4 |
| Participant selection | | | | |
| 10. | Sampling | How were participants selected? *e.g. purposive, convenience, consecutive, snowball* | We approached 129 clinical/institutional contacts in order to recruit interview partners. Our recruiting strategy was driven by theoretical sampling approach and we aimed to include different clinical settings in Germany. | page 3 |
| 11. | Method of approach | How were participants approached? e*.g. face-to-face, telephone, mail, email* | E-Mail, telephone, social media, flyer | page 3 |
| 12. | Sample size | How many participants were in the study? | 39 | page 3 |
| 13. | Non-participation | How many people refused to participate or dropped out? Reasons? | There was no drop-out.<br>Two contacted individuals actively declined to participate in the study at the first point of contact without specific reasons, while two asked to be contacted only after the pandemic. | page 3 |
| Setting | | | | |
| 14. | Setting of data collection | Where was the data collected? e*.g. home, clinic, workplace* | Interviews were conducted by telephone or via a suitable data protection compliant video conferencing system. | page 3 |
| 15. | Presence of non-participants | Was anyone else present besides the participants and researchers? | No. | |
| 16. | Description of sample | What are the important characteristics of the sample? *e.g. demographic data, date* | Relevant demographic data are shown in Figure 1. | page 5 |
| Data collection | | | | |

| No | Item | Guide questions/description | Answers | Referred in manuscript on |
|---|---|---|---|---|
| 17. | Interview guide | Were questions, prompts, guides provided by the authors? Was it pilot tested? | A topic guide was developed and used where appropriate. As the question was "I would like to ask you to tell us how you experience the phase of preparation for the care of COVID-19 infected patients.", this phrase encouraged the interviewees to talk as much freely as possible.<br>Two interviews were determined as pilot interviews and included in the sample.<br>The topic guide is attached as supplementary information. | page 4 |
| 18. | Repeat interviews | Were repeat interviews carried out? If yes, how many? | Repeat interviews are carried out and will be published at a later stage. | page 3 |
| 19. | Audio/visual recording | Did the research use audio or visual recording to collect the data? | Two interviews were conducted including visual recording, 37 with audio only. | page 3 |
| 20. | Field notes | Were field notes made during and/or after the interview or focus group? | Yes, we documented the results of the interpretation groups in tabular form. | page 4 |
| 21. | Duration | What was the duration of the interviews or focus group? | see Figure 1 | page 5 |
| 22. | Data saturation | Was data saturation discussed? | In our continuous interpretation groups, saturation was discussed once per week. | |
| 23. | Transcripts returned | Were transcripts returned to participants for comment and/or correction? | No. | page 4 |
| **Domain 3: analysis and findings** | | | | |
| Data analysis | | | | |
| 24. | Number of data coders | How many data coders coded the data? | Ten | page 4 |
| 25. | Description of the coding tree | Did authors provide a description of the coding tree? | The inductively created concepts and categories are our coding tree. | page 5-6 (Categories and concepts) |
| 26. | Derivation of themes | Were themes identified in advance or derived from the data? | Yes, we identified seven major categories and 22 concepts from our data, see figure 2. | page 8, particular figure 2 |

| No | Item | Guide questions/description | Answers | Referred in manuscript on |
|---|---|---|---|---|
| 27. | Software | What software, if applicable, was used to manage the data? | Interview data was managed with MAXQDA by Verbi GmbH or ATLAS.ti by Scientific Software Development GmbH.<br>For collaborative interpretation, we used Confluence by Atlassian (v. 7.5.2). | page 4 |
| 28. | Participant checking | Did participants provide feedback on the findings? | We did not consider respondent validation. | page 4 |
| Reporting | | | | |
| 29. | Quotations presented | Were participant quotations presented to illustrate the themes / findings? Was each quotation identified? e*.g. participant number* | Quotations can be found in Table 1. The quotations can be identified by our generated named pseudonyms. Remark: These pseudonyms do not allow any conclusions to be drawn about the true identity of the study participants. | page 9-10 |
| 30. | Data and findings consistent | Was there consistency between the data presented and the findings? | Yes. We transparently documented all interpretation process steps and see consistency after all. | page 4 |
| 31. | Clarity of major themes | Were major themes clearly presented in the findings? | We identified and clearly presented seven major themes in the results section. | page 5 |
| 32. | Clarity of minor themes | Is there a description of diverse cases or discussion of minor themes? | We aimed to illustrate the heterogeneity of the topics of our data as comprehensively as possible. We found that the identified categories and concepts in our data were highly dynamic. | page 12 |
